# Overlap between ultra-processed food and food that is high in fat, salt or sugar: analysis of 11 annual waves of the UK National Diet and Nutrition Survey 2008/09-2018/19

**DOI:** 10.1101/2024.08.27.24312650

**Authors:** Viktorija Kesaite, Yanaina Chavez-Ugalde, Martin White, Jean Adams

## Abstract

While many countries use guidance and policies based on nutrients and food groups to support citizens to consume healthy diets, fewer have explicitly adopted the concept of ultra-processed foods (UPF). UPF consumption is associated with many adverse health outcomes in cohort studies. In the UK, a nutrient profiling model (NPM) is used to identify foods high in fat, salt or sugar (HFSS) and several policies target these. It is not known how well the NPM also captures UPF. We aimed to quantify the proportion of food and drink items consumed in the UK that are HFSS, UPF, both or neither and describe the food groups making the largest contributions to each category. We analysed data from the National Diet and Nutrition Survey (NDNS), between 2008/09 and 2018/19, using descriptive statistics. We used three metrics of food consumption: all foods, % of energy in all foods (reflecting that different foods are consumed in different portion sizes and are of different energy densities), and % of food weight in all foods (reflecting that some UPFs have few calories but are consumed in large volumes). We found that, 33.4% of foods, 47.4% of energy, and 16.0% of food weight were HFSS; 36.2%, 59.8% and 32.9% respectively were UPFs; 20.1%, 35.1% and 12.6% were both; and 50.5%, 27.9% and 63.7% were neither. In total, 55.6% of UPF foods, 58.7% of energy from UPFs and 38.3% of food weight from UPF consumed were also HFSS. The most common food groups contributing to foods that were UPF but not HFSS were low calorie soft drinks and white bread. The UK NPM captures at best just over half of UPFs consumed in the UK. Expanding the NPM to include ingredients common in UPFs would capture a larger percentage of UPFs and could incentivise “de-formulation” of UPF products.

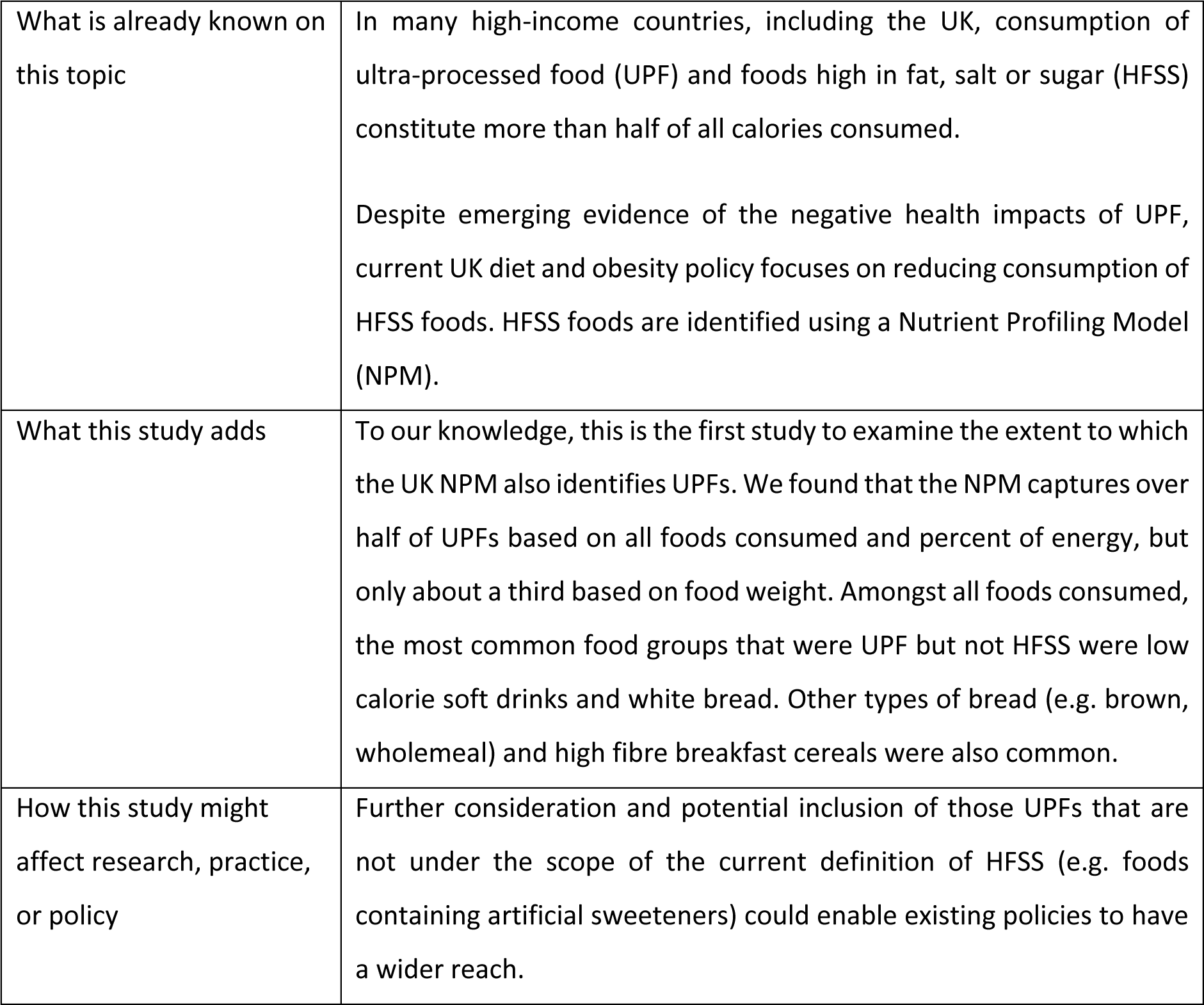

|  |  |
| --- | --- |
| What is already known on this topic | In many high-income countries, including the UK, consumption of ultra-processed food (UPF) and foods high in fat, salt or sugar (HFSS) constitute more than half of all calories consumed.<br>Despite emerging evidence of the negative health impacts of UPF, current UK diet and obesity policy focuses on reducing consumption of HFSS foods. HFSS foods are identified using a Nutrient Profiling Model (NPM). |
| What this study adds | To our knowledge, this is the first study to examine the extent to which the UK NPM also identifies UPFs. We found that the NPM captures over half of UPFs based on all foods consumed and percent of energy, but only about a third based on food weight. Amongst all foods consumed, the most common food groups that were UPF but not HFSS were low calorie soft drinks and white bread. Other types of bread (e.g. brown, wholemeal) and high fibre breakfast cereals were also common. |
| How this study might affect research, practice, or policy | Further consideration and potential inclusion of those UPFs that are not under the scope of the current definition of HFSS (e.g. foods containing artificial sweeteners) could enable existing policies to have a wider reach. |

## INTRODUCTION

Numerous studies have shown that diets high in some components (e.g. sodium, sugar, fat and calories) and low in others (e.g. whole grains and fruits) increase the risk of non-communicable diseases such as cardiovascular disease and cancer (1). Recent estimates suggest that food and drinks (hereafter: ‘foods’) high in fat, salt, or sugar (HFSS) constitute more than half of total energy intake among adults in the UK (2, 3). Similar estimates are observed in the consumption of ultra-processed foods (UPF) (4).

In the UK, a Nutrient Profiling Model (NPM) is used to identify HFSS foods and restrict advertising on television (5), out-of-doors in some local authorities and where foods can be placed in grocery stores (6). The NPM was originally developed in 2004/05, and defines foods as being HFSS based on energy, saturated fat, total sugar, sodium, fibre, protein, and fruit, vegetable, and nut content per 100g (7). An updated version was developed in 2018 (8), based on revised guidance on sugar and fibre intake (9) however, despite consultations, has not yet been adopted into practice (10).

High consumption of ultra-processed foods (UPF), as identified by the Nova classification (11), has also been negatively associated with adverse health outcomes (12). The Nova system is the most common method of classifying foods based on their degree of food processing used in the academic literature and has been identified as the most applicable to the UK (10). Recent Euromonitor statistics suggest that the highest volume of sales of UPFs globally are in Western Europe, North America, and Australasia, with increasing trends observed elsewhere (13, 14). Findings based on a systematic review indicate that the US and the UK had the highest proportion of energy intake from UPFs (>50%)(4).

The UK’s approach to public health nutrition, where nutrients and food groups such as fruit and vegetables, are used to drive guidance and policy reflects similar approaches in many countries. However, a growing number of countries have now also included specific reference to avoidance of UPFs in their dietary guidance (15).

As policy makers contend with whether to focus policy and dietary guidance on UPFs, an important question is: how well do current approaches, such as the UK’s NPM, also identify UPFs? If most UPFs are also HFSS then the benefits of introducing additional UPF-focused policy and guidance may be minimal. We are aware of only one other study examining the overlap between UPFs and other methods of identifying less-healthy foods (3). This recent US study found that around three-quarters of UPF foods purchased were also HFSS and that extending the HFSS definition to include non-nutritive sweeteners, flavourings, colours and additives would capture nearly 100% of UPFs (3). To the best of our knowledge no previous work has explored the overlap between HFSS, as identified by the UK NMP, and UPF in the UK.

In this study, our aim was to quantify the proportion of foods consumed in the UK that are HFSS, UPF, both or neither.

## METHODS

We used data from eleven waves (2008/9 – 2018/19) of the UK National Diet and Nutrition Survey (NDNS).

### Data description

NDNS is a continuous cross-sectional survey conducted every year in the UK. It collects data on food consumption, nutrient intake, and nutritional status of the general population aged 1.5 years and over residing in private UK households. Each year, a multistage probability design is used to generate a new random sample. Selected household addresses are clustered into small geographical units called Primary Sampling Units. Households are randomly selected from these units, and participants are randomly selected from each household. Full data is collected from about 500 children and 500 adults each year.

In 2008-19, all NDNS participants self-reported their food and beverage consumption over three or four days using a written food diary, with portion sizes recorded using standard household measures and product labels. Parents and guardians reported on behalf of children under 11 years. Food diary data are entered into the Diets In Nutrients Out (DINO) database and food codes are used to link foods to the NDNS nutrient composition database, which includes nutritional information on more than 6000 foods (16). The response rate to food diaries is 50% or more. The NDNS provides sample weights to reduce the risk of non-random selection (16).

### Categorisation of foods as HFSS and UPF

We categorised foods as HFSS or not based on the UK NPM 2004/5 and, in a sensitivity analysis, using the 2018 NPM. We used the Nova framework to classify foods as UPF or not, using assignments from previous work (17) (18). In this, two researchers independently classified foods into the four Nova groups. To assign Nova group, NDNS foods were coded where possible based on their NDNS main food group (n=56). If there was uncertainty about whether all foods in a main food group would have the same Nova category, foods were categorised where possible by food subgroups (n=136). In cases of further uncertainty, individual food items (n=4,555) were classified (e.g., composite dishes which include individual foods from more than one main or sub-group). Any disagreement between the researchers on the classifications were resolved through discussion. This process achieved a high degree of inter-rater reliability - 97% in the first and 99.8% in the second round after discussion (19). Food supplements (i.e., vitamins and minerals) are not classified by the Nova system and we did not include them or alcoholic beverages.

### Data analyses

Reflecting our aims, our analyses were descriptive. Firstly, we conducted a food-level analysis. This used all reported foods consumed by NDNS participants in 2008/09-2018/19 with foods reported multiple times being included multiple times, which effectively weights the findings for consumption. We described the percentage of foods that were UPF, HFSS, neither or both. We present this first as a food level analyses including all foods and, separately, the relative contribution of UPFs and HFSS to total food energy (kcal) and total food weight (g) intake. We then present examples of food groups making the greatest contributions to each category based on all foods, total food energy (kcal), and total weight (g), and finally present a person-day-level analysis which replicates the food level analysis adjusting for non-response bias. We include an analysis based on total food energy to reflect that different foods are consumed in different portion sizes and have different energy densities and so make different relative contributions to the diet. This enables comparisons with other studies (3). We also include a metric based on total food weight to reflect that some UPFs make little contribution to energy (e.g., artificially sweetened beverages), and list the NDNS main food groups making the largest contributions to each of these four categories (UPF, HFSS, both and neither).

Secondly, we conducted person-level analyses. These replicate the food level analyses but by including population survey weights we additionally account for non-response bias. As a robustness check, we replicated the food-level analyses of foods consumed for men and women separately. The study protocol is available at https://osf.io/hr6f3/, the code and the libraries for calculating HFSS scores and the analyses is available at https://github.com/VKesaite/HFSS-and-UPF. All calculations were carried out in Python version 3.11.4. There were no substantive changes to the protocol.

## RESULTS

A total of 15,655 individuals (7,207 males and 8,448 females) were included in the analysis. They reported consuming 1,730,158 foods, representing 4,555 unique food names. We report findings based on the 2004/05 NPM here and on the 2018 NPM in supplemental material.

### Food-level analysis

Figure 1 shows Venn diagrams describing the proportions of (a) foods, (b) food energy and (c) food weight that were HFSS (right inner circle), UPF (left inner circle), both (inner circle overlap) and neither (outer circle). The proportion of UPFs that are HFSS is also shown. In total, 33.4% of foods, 47.4% of energy and 16.0% of food weight was HFSS, whilst 36.2% of food, 59.8% of energy and 32.9% of food weight was UPF. We found that 55.6% of UPFs, 58.7% of energy from UPFs, and 38.3% of UPF weight was also HFSS. Robustness checks in males and females separately showed similar findings (Table S1). Analysis using the 2018 version of the NPM led to broadly similar results, but in general a slightly smaller proportion of UPF was categorised as HFSS compared to the 2004/05 NPM (Table S1).

**Figure 1.**
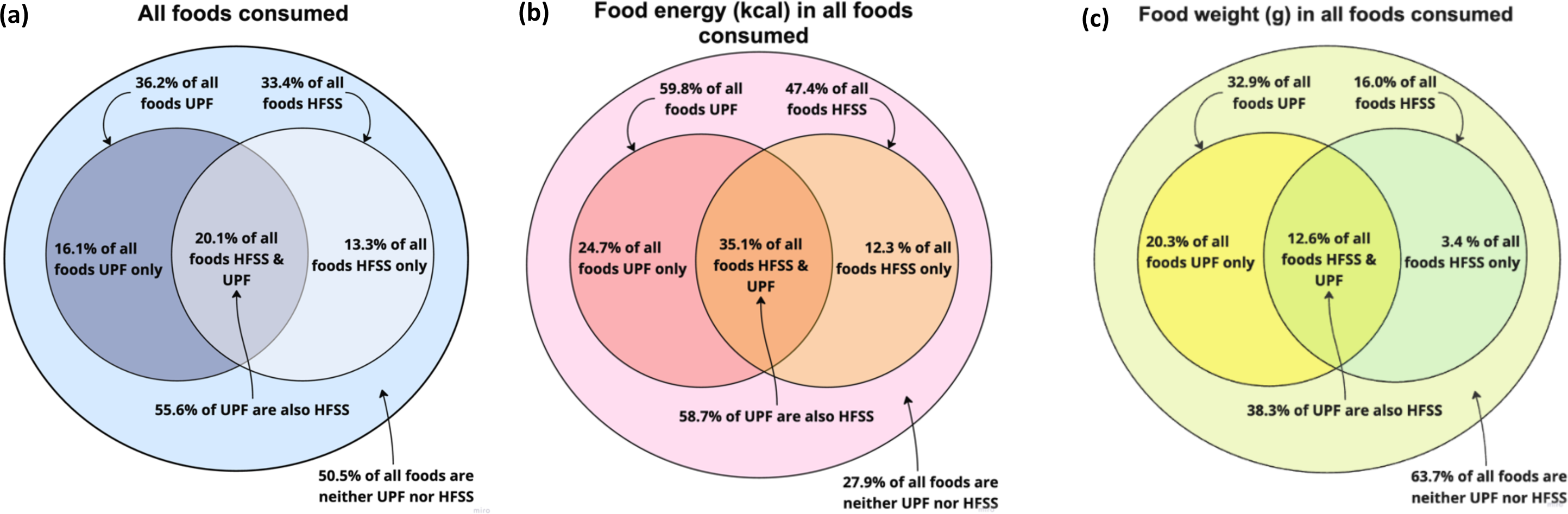
The percentage of (a) all foods, (b) food energy in kcal, and (c) food weight in grams consumed that was derived from foods that are high in fat, salt or sugar, ultra-processed foods, both or neither; UK National Diet and Nutrition Survey 2008/9-2018 *Note.* HFSS = high in fat, salt or sugar (using the 2004/05 Nutrient Profiling Model); UPF = ultra-processed food

Table 1 lists the ten main food groups making the greatest contributions to the categories HFSS only, UPF only, both and neither for all foods. Tables 2 and 3 show similar data for food energy and food weight respectively. The most common main food groups contributing to foods and food energy that were HFSS but not UPF reflected products high in sugars (i.e. ‘sugars, preserves and sweet spread’) and dairy products (or replacements) such as cheese, butter, oil, butter and oil replacements and whole milk. When analysed by food weight, the most common food groups contributing to the HFSS only category were dairy products such as whole milk, cheese, butter, and cream products, and also sugars (i.e. ‘sugars, preserves and sweet spread’), bacon and ham products, and starchy products (i.e. ‘pasta, rice & other cereals’).

**Table 1.** The ten main food groups (% contribution) making the greatest contributions to the categories: high in fat, salt or sugar only, ultra-processed food only, both and neither at the food level for all foods; UK National Diet and Nutrition Survey 2008/9-2018/19

| All foods consumed |  |  |  |
| --- | --- | --- | --- |
| HFSS only | UPF only | HFSS and UPF | Neither HFSS nor UPF |
| Sugars, preserves & sweet spread (26.6%) | Soft drinks, low calorie (21.3%) | Miscellaneous* (14.0%) | Tea, coffee & water (31.0%) |
| Miscellaneous* (13.1%) | White bread (17.5%) | Biscuits (10.1%) | Vegetables not raw (13.0%) |
| Cheese (10.9%) | Miscellaneous* (5.9%) | Reduced fat spread (9.6%) | Semi-skimmed milk (12.3%) |
| Butter (10.4%) | Chips, fried & roast potatoes, potato products (5.8%) | Soft drinks, not low calorie (8.2%) | Fruit** (9.8%) |
| Other margarine, fats & oils† (9.2%) | High fibre breakfast cereals (5.5%) | Crisps & savoury snacks (6.1%) | Salad & other raw vegetables (7.9%) |
| Whole milk (7.9%) | Brown, granary & wheatgerm bread (5.5%) | Chocolate confectionery (6.1%) | Pasta, rice & other cereals (4.3%) |
| Bacon & ham (4.1%) | Yoghurt, fromage, frais, & dairy desserts (5.4%) | Buns, cakes, pastries & fruit pies (5.4%) | Miscellaneous* (3.4%) |
| Pufa margarine & oils (3.8%) | Wholemeal bread (5.3%) | Bacon & ham (3.6%) | Other potatoes, potato salads & dishes** (2.6%) |
| Nuts & seeds (2.9%) | Vegetables not raw (4.0%) | Other breakfast cereals** (3.5%) | Fruit juice (2.6%) |
| Other milk & cream† (2.7%) | Artificial sweeteners (3.5%) | Sugars, preserves & sweet spreads (3.5%) | Chicken & turkey dishes (2.3%) |

*Note:* HFSS = high in fat, salt or sugar (using the 2004/05 Nutrient Profiling Model); UPF = ultra-processed food ; main food groups are those used in the National Diet and Nutrition Survey; \*Miscellaneous includes e.g. mayonnaise, sour cream dip, soups; \*\*Fruit includes e.g. raw fruit, fruit stewed with and without sugar, fruit canned in fruit and sugar syrup; †Other milk & cream includes e.g. plant-based milk alternatives, coffee creamer, crème fraîche; ††Other breakfast cereals includes e.g. special flakes, rice crispies; †Other margarine, fats & oils includes e.g. vegetable suet, palm oil, chicken fat; ††Other potatoes, potato salads & dishes includes potato curry, mash potato with butter and cream.

**Table 2.** The ten main food groups (% contribution) making the greatest contributions to the categories: high in fat, salt or sugar only, ultra-processed food only, both and neither at the food level for food energy (kcal); UK National Diet and Nutrition Survey 2008/9-2018/19

| Food energy (kcal) in all foods consumed |  |  |  |
| --- | --- | --- | --- |
| HFSS only | UPF only | HFSS and UPF | Neither HFSS nor UPF |
| Cheese (22.2%) | White bread (27.8%) | Biscuits (11.2%) | Pasta, rice & other cereals (16.2%) |
| Butter (14.6%) | Chips fried & roast potatoes & potato products (12.7%) | Buns, cakes, pastries & fruit pies (10.0%) | Fruit** (11.9%) |
| Sugars, preserves & sweet spreads (12.9%) | Brown granary & wheatgerm bread (8.4%) | Chocolate confectionery (7.8%) | Semi skimmed milk (10.8%) |
| Whole milk (8.6%) | Wholemeal bread (7.3%) | Crisps & savoury snacks (7.2%) | Chicken & turkey dishes (9.3%) |
| Bacon and ham (8.2%) | High fibre breakfast cereals (6.9%) | Pasta, rice & other cereals (7.1%) | Other potatoes, potato salads & dishes** (7.7%) |
| Other margarine, fats & oils<sup>†</sup> (6.0%) | Pasta, rice & other cereals (5.5%) | Soft drinks, not low calorie (7.0%) | Vegetables not raw (7.0%) |
| Nuts & seeds (4.3%) | Vegetables not raw (4.3%) | Miscellaneous* (5.8%) | Beef, veal & dishes (5.5%) |
| Other milk & cream<sup>†</sup> (3.0%) | Yoghurt, fromage, frais & dairy desserts (3.9%) | Sausages (5.4%) | Eggs, and egg dishes (4.7%) |
| Buns, cakes, pastries (2.2%) | Miscellaneous* (3.5%) | Reduced fat spread (5.1%) | Fruit juice (4.3%) |
| Pufa margarine & oils (2.1%) | Coated chicken (3.3%) | Meat pies & pastries (4.1%) | Chips fried & roast potatoes, and potato products (4.1%) |
| Pasta, rice & other cereals (2.0%) | Other milk & cream<sup>†</sup> (1.9%) | Other breakfast cereals (3.8%) | Whole milk (4.0%) |

*Note:* HFSS = high in fat, salt or sugar (using the 2004/05 Nutrient Profiling Model); UPF = ultra-processed food ; main food groups are those used in the National Diet and Nutrition Survey; \*Miscellaneous includes e.g. mayonnaise, sour cream dip, soups; \*\*Fruit includes e.g. raw fruit, fruit stewed with and without sugar, fruit canned in fruit and sugar syrup; <sup>†</sup>Other milk & cream includes e.g. plant-based milk alternatives, coffee creamer, crème fraiche; <sup>††</sup>Other breakfast cereals includes e.g. special flakes, rice crispies; <sup>†</sup>Other margarine, fats & oils includes e.g. vegetable suet, palm oil, chicken fat; <sup>††</sup>Other potatoes, potato salads & dishes includes potato curry, mash potato with butter and cream.

**Table 3.** The ten main food groups (% contribution) making the greatest contributions to the categories: high in fat, salt or sugar only, ultra-processed food only, both and neither at the food level for food weight (g); UK National Diet and Nutrition Survey 2008/9-2018/19

| Food weight (g) in all foods consumed |  |  |  |
| --- | --- | --- | --- |
| HFSS only | UPF only | HFSS and UPF | Neither HFSS nor UPF |
| Whole milk (34.4%) | Soft drinks, low calorie (35.4%) | Soft drinks, not low calorie (34.3%) | Tea, coffee & water (57.3%) |
| Cheese (15.2%) | White bread (9.9%) | Pasta, rice & other cereals (6.2%) | Semi-skimmed milk (7.9%) |
| Sugars, preserves, & sweet spreads (9.3%) | Soft drinks, not low calorie (6.5%) | Miscellaneous* (5.7%) | Fruit** (6.7%) |
| Bacon and ham (9.1%) | Miscellaneous* (6.1%) | Buns, cakes, pastries & fruit pies (5.6%) | Vegetables, not raw (5.6%) |
| Butter (5.4%) | Chips fried & roast potatoes, & potato products (5.5%) | Biscuits (5.1%) | Fruit juice (3.7%) |
| Other milk & cream<sup>†</sup> (2.4%) | Yoghurt, fromage, frais & dairy desserts (4.7%) | Sausages (4.3%) | Pasta, rice, & other cereals (3.3%) |
| Pasta, rice & other cereals (2.0%) | Vegetables not raw (4.3%) | Yoghurt, fromage, frais & dairy desserts (3.4%) | Other potatoes, potato salads & dishes<sup>††</sup> (3.1%) |
| Nuts & seeds (2.0%) | Tea, coffee & water (3.4%) | Chocolate confectionery (3.3%) | Whole milk (2.1%) |
| Other margarine, fats & oils<sup>‡</sup> (1.8%) | Pasta, rice & other cereals (3.4%) | Crisps & savoury snacks (3.0%) | Salad & other raw vegetables (2.0%) |
| White fish coated or fried (1.7%) | Brown granary & wheatgerm bread (3.1%) | Ice cream (2.9%) | Chicken & turkey dishes (1.9%) |

*Note:* HFSS = high in fat, salt or sugar (using the 2004/05 Nutrient Profiling Model); UPF = ultra-processed food ; main food groups are those used in the National Diet and Nutrition Survey; \*Miscellaneous includes e.g. mayonnaise, sour cream dip, soups; \*\*Fruit includes e.g. raw fruit, fruit stewed with and without sugar, fruit canned in fruit and sugar syrup; <sup>†</sup>Other milk & cream includes e.g. plant-based milk alternatives, coffee creamer, crème fraîche; <sup>‡</sup>Other margarine, fats & oils includes e.g. vegetable suet, palm oil, chicken fat; <sup>††</sup>Other potatoes, potato salads & dishes includes potato curry, mash potato with butter and cream.

The most common main food groups contributing to foods in the category UPF but not HFSS often included non-nutritive sweeteners (i.e. ‘soft drinks low calorie’, ‘artificial sweeteners’) or were breads including white, brown, granary and wholemeal bread. Fried and roast potatoes and potato products were also a common food group in this category. When considering food weight and food energy, the most common food groups in the UPF only category were similar to total food category.

In the category of foods that were UPF and HFSS, the most common main groups reflected miscellaneous foods (e.g., condiments and soups), bakery products (e.g. biscuits and ‘buns, cakes, pastries & fruit pies’), reduced fat spread, and ‘soft drinks, not low calorie’. When considering by food energy and food weight, products high in sugar (biscuits, ‘buns, cakes, pastries & fruit pies’, chocolate confectionery), ‘crisps and savoury snacks’, and ‘pasta, rice & other cereals’ made substantial contributions to the category that was both UPF and HFSS.

The most common main food group contributing to the category of foods that were neither HFSS nor UPF was tea, coffee and water. Cooked and raw vegetables, semi-skimmed milk and fruit were also were also prominent. Similar findings were seen for food weight. By food energy, the food groups making the largest contribution to this category included pasta, rice & other cereals, fruit, semi-skimmed milk, and chicken and turkey dishes.

### Person-level analysis

The results of the person-level analysis are summarised in Table 4. Additionally adjusting for non-response bias led to small changes compared to the estimates in Figure 1. At person-level, 55.1% of UPF foods are also HFSS (c.f. 55.6% at food level), 53.8% of UPF energy is also HFSS (c.f. 58.7%), and 40.0% of UPF food weight is also HFSS (c.f. 38.3%).

**Table 4.** The percentage of per capita daily foods, food energy in kcal, and food weight in grams consumed that was derived from foods that are high in fat, salt or sugar, ultra-processed foods, both and neither; UK National Diet and Nutrition Survey 2008/9-2018/19

|  | All foods consumed; % (SD) | Food energy (kcal); % (SD) | Food weight (g); % (SD) |
| --- | --- | --- | --- |
| Neither UPFs nor HFSS | 52.9 (14.4) | 31.7 (13.6) | 66.8 (18.6) |
| All UPF | 33.6 (15.6) | 54.9 (15.7) | 29.7 (18.3) |
| UPF only | 15.0 (8.9) | 24.6 (10.5) | 18.2 (13.4) |
| All HFSS | 32.0 (10.6) | 43.7 (13.2) | 15.0 (10.9) |
| HFSS only | 13.5 (8.1) | 13.4 (8.7) | 3.5 (4.3) |
| HFSS and UPFs | 18.5 (10.0) | 30.3 (13.8) | 11.5 (10.0) |
| UPF that is also HFSS | 55.1 (15.5) | 53.9 (17.3) | 40.0 (19.0) |

*Note:* HFSS = high in fat, salt or sugar (using the 2004/05 Nutrient Profiling Model); UPF = ultra-processed food; data are weighted to correct for non-random non-response of households.

Analysis using the 2018 version of the NPM led to broadly similar results, but overall a slightly smaller proportion of UPF was categorised as HFSS compared to the 2004/05 NPM (Table S2).

## DISCUSSION

### Summary of main findings

This is the first study to assess the extent to which foods consumed in the UK are HFSS, UPFs, both or neither. Using 11 years of data from NDNS we found that around one third of all foods consumed (33.4%) were HFSS, one third (36.2%) were UPF, one fifth were both (20.1%) and half (50.5%) neither. We found that 55.6% of UPFs, 58.7% of food energy and 38.3% of food weight consumed were also HFSS. This means that policies focused on HFSS reduction also target between one third and just over a half of UPFs.

When considering all foods consumed, common foods that were UPF but not HFSS (and so would not be included in HFSS-focused policies) included low calorie soft drinks, manufactured breads, and high fibre breakfast cereals. Food groups making substantial contributions to all foods consumed in the category HFSS and UPF were miscellaneous (e.g., condiments and soups), biscuits, reduced fat spread, and ‘soft drinks, not low calorie’.

### Strengths and limitations

To the best of our knowledge, this is the first study to classify all foods consumed in the UK as both HFSS and UPF and compare these classifications. We used food-level dietary data, with 3 or 4 food diary days per person from the NDNS (20, 21). Food diaries reduce the recall bias common in food frequency questionnaires and 24-hour recalls (21, 22). We used three metrics of food consumption – total number of foods, food energy, and food weight. Analysing food energy recognises that different foods are consumed in different portion sizes while assessing food weight identifies, in particular, foods that have a high weight (e.g. drinks) but low energy ingredients (e.g. non-nutritive sweeteners).

Whilst the accuracy of food classification using the Nova system has been criticised as poorly replicable (23), we and others have achieved a high degree of inter-rater reliability (19, 24). Furthermore, by including all foods reported by NDNS participants (excluding alcohol and supplements), we effectively weighted by frequency of consumption. Additional adjustment for non-response bias by including survey weights in individual-level analyses did not change the overall pattern of findings indicating that our findings are likely to be generalisable to the UK population.

Despite their strengths, food diaries are self-reported leading to the potential for measurement error. Further, whilst we included 11 years of data, the most recent was from 2019. Given the increasing number of new foods commercially available (25), we may not have captured recent changes in food consumption. Moreover, combining 11 years of data might have masked important changes over time, but ensured a large sample size. NDNS did not collect data from new participants in 2020 and subsequently changed dietary data collection method meaning that newer comparable data is not available.

There are also some potential limitations with the nutritional measures used in this study. The Nova classification has been criticised for its lack of consideration of nutritional content and quality. With some foods, ultra-processing might yield nutritional and health benefits such as micronutrient fortification, increased affordability, extended shelf life and reduced waste (26). A recent study indicated that while consumption of some UPFs, such as artificially and sugar-sweetened beverages, was associated with an increased risk of cancer and cardiometabolic diseases, no evidence of an association was found for others such as UPF bread and cereals (1). Our second measure of nutritional quality, HFSS has also been criticised as an oversimplified measure of nutritional quality (27).

### Interpretation and implications

We found that, at best, 58.7% of UPFs consumed over 11 years of the UK NDNS were identified as HFSS. This is a lower degree of overlap than a recent US study which found that 75.4% of UPF were also HFSS (3). These differences may reflect the different food supply, purchase and consumption patterns seen in the UK and USA.

Our findings suggests that current UK policy which focuses on HFSS captures at best just over half of UPFs, but some key UPFs are not included. Amongst all foods consumed, common food groups that were UPF but not HFSS included ‘low calorie soft drinks’, bread, and miscellaneous products (e.g., condiments and soups).

Extending the definition of HFSS to include non-nutritive sweeteners would increase the proportion of UPF captured by HFSS to just over two-thirds. Similar findings were reported by Popkin et al (2024) (3). Whilst some recent UK public health nutrition policies have increased the number of products available that contain non-nutritive sweeteners (28), the World Health Organization has recently noted that these may not be useful for weight loss (29). There may also be other ways that the HFSS algorithm could be adapted to capture more UPFs. However, there remains a lack of consensus among policymakers and scientists on whether the evidence on the health harms of UPF is strong enough to justify regulation (10).

When analysing by food energy, rather than total number of foods the ‘other milk & cream’ food group was among the top ten food groups contributing to the category of UPF but not HFSS. This includes plant-based milk alternatives. Some plant-based alternatives can be healthier than equivalent meat and dairy products (30) and some plant-based alternatives are UPF. Whilst there is uncertainty about the environmental impacts of UPF as a category (31), reducing consumption of animal-based products at a population level will be essential to achieve net-zero (32). UPF plant-based alternatives could be a stepping stone to less processed plant-based alternatives (33, 34). This highlights the trade-offs common in individual dietary choices as well as policymaking and why simply dichotomising foods into those that should be consumed or avoided is challenging, however that dichotomisation is achieved. Nevertheless, food profiling methods that consider both human and planetary health would allow for a more holistic approach, and regulating foods on sustainability grounds is likely to have health-co-benefits (35).

We found that many food groups are common contributors to both HFSS only, UPF only, both and neither. For example, regardless of the metric of consumption (foods, food energy or food weight), the main food group ‘miscellaneous’ is in the top ten contributors to many categories. This reflects that not all foods within main food groups necessarily share the same UPF or HFSS status. For example, homemade soups could be HFSS whilst manufactured soups are likely to be UPF and may be HFSS.

Much policy in the UK has focused on food reformulation, whereby manufacturers are incentivised to change the composition of foods to ensure they are not HFSS (36). Other policies have focused specifically on reducing sugar and calorie content (37). Whilst there is evidence that some of these policies have prompted reformulation (e.g. removal of sugar from soft drinks) (38) and have been associated with overall reduction in sugar consumption(39) and health benefits (40), reformulated foods which are no longer HFSS often remain UPF (e.g. artificially sweetened soft drinks). An alternative approach using policies that incentivise de-formulation (41) (i.e. removing components and additives commonly used in UPFs, such as non-nutritive sweeteners) could help reduce production and consumption of UPF. Accompanying such policy approaches with those that incentivise whole food consumption could offer important synergies.

The difference in results we find by analysing the data by all foods, food energy and food weight reflect differences in the food energy and weight of different foods. For example, low calorie soft drinks make up a substantial proportion of UPF only foods and food weight, but not food energy. This reflects that these products are low in energy but may be consumed in large portions.

The 2018 NPM was developed to take account of updated guidance on sugar and fibre consumption (9). Whilst this generated broadly similar results to using the 2004/05 NPM, in most cases a slightly smaller proportion of UPF were categorised as HFSS (e.g. 55.6% of UPF foods consumed were HFSS using the 2004/05 NPM and 54.4% using the 2018 NPM). These small differences are unlikely to be of policy relevance.

Our results reinforce the well-established finding that current UK diets are sub-optimal for health. After adjustment for non-response bias, we found that 54.9% of per capita daily energy was derived from UPFs and 43.7% from HFSS foods. This is comparable to previous findings. For instance, a previous study in the UK based on NDNS data from 2008-14 found that 56.8% of energy was derived from UPFs (42). Similarly, in Canada 47.7% of energy was derived from UPFs (43), and in the US 57.5% was derived from UPFs (44). Studies from other countries find a smaller proportion of energy attributable to UPFs. For example, a study from Italy found that 17.3% of energy was derived from UPFs (45). Current UK dietary guidance advises citizens to eat HFSS foods “less often and in small amounts” (46).

## CONCLUSION

In this analysis of 11 years of data from the UK NDNS, we found that the NPM, used to identify foods that are HFSS for regulatory purposes identifies, at best, 58.7% of UPFs. If UK policymakers decide that regulation of UPFs is necessary, additional action will be required to extend current policy. This could involve extending the current NPM to include ingredients, such as non-nutritive sweeteners, that are common in foods that are UPF but not currently identified as HFSS. Further work is required to confirm that UPF consumption is causally associated with health harms and to determine the environmental impacts of UPF.

## Data Availability

All data produced are available online at UK DATA SERVICE
https://beta.ukdataservice.ac.uk/datacatalogue/series/series?id=2000033

## ACKNOWLEDGEMENTS

We are grateful to Birdem Amoutzopoulos and David Collins for reviewing the code for categorising foods high in fat, salt or sugar.

## RESEARCH ETHICS APPROVAL

No ethics approval was required.

