## Appendix Tables for "Overlap between ultra-processed food and food that is high in fat, salt or sugar: analysis of 11 annual waves of the UK National Diet and Nutrition Survey 2008/09-2018/19"

**Table S1. The percentage of all foods, food energy in kcal, and food weight in grams consumed that was derived from foods that are high in fat, salt or sugar, ultra-processed foods, both and neither; UK National Diet and Nutrition Survey 2008/9-2018/19**

| | All foods consumed; % | | | Food energy (kcal); % | | | Food weight (g); % | | |
|---|---|---|---|---|---|---|---|---|---|
| | All participants | Male participants | Female participants | All participants | Male participants | Female participants | All participants | Male participants | Female participants |
| **Neither UPFs nor HFSS (NPM 2004/5)** | 50.5 | 48.2 | 52.4 | 27.9 | 26.8 | 29.0 | 63.7 | 60.7 | 66.4 |
| **Neither UPFs nor HFSS (NPM 2018)** | 51.7 | 49.4 | 53.6 | 28.9 | 27.9 | 30.0 | 62.8 | 59.8 | 65.5 |
| **All UPF** | 36.2 | 38.2 | 34.6 | 59.8 | 61.0 | 58.6 | 32.9 | 35.6 | 30.5 |
| **UPF only (NPM 2004/5)** | 16.1 | 16.9 | 15.3 | 24.7 | 25.3 | 24.1 | 20.3 | 21.5 | 19.2 |
| **UPF only (NPM 2018)** | 16.5 | 17.3 | 15.8 | 27.3 | 28.1 | 26.6 | 19.5 | 20.7 | 18.4 |
| **All HFSS (NPM 2004/5)** | 33.4 | 34.8 | 32.2 | 47.4 | 47.9 | 46.9 | 16.0 | 17.8 | 14.4 |
| **All HFSS (NPM 2018)** | 31.8 | 33.2 | 30.6 | 43.7 | 44.0 | 43.5 | 17.7 | 19.5 | 16.1 |
| **HFSS only (NPM 2004/5)** | 13.3 | 13.6 | 13.0 | 12.3 | 12.2 | 12.3 | 3.4 | 3.8 | 3.5 |
| **HFSS only (NPM 2018)** | 12.1 | 12.4 | 11.8 | 11.3 | 11.1 | 11.4 | 4.3 | 4.6 | 4.6 |
| **HFSS and UPF (NPM 2004/05)** | 20.1 | 21.2 | 19.2 | 35.1 | 35.7 | 34.5 | 12.6 | 14.1 | 11.3 |
| **HFSS and UPF (NPM 2018)** | 19.7 | 20.8 | 18.8 | 32.5 | 32.9 | 32.0 | 13.4 | 14.8 | 12.1 |
| **UPF that is also HFSS (NPM 2004/5)** | 55.6 | 55.6 | 55.6 | 58.7 | 58.5 | 58.9 | 38.3 | 39.5 | 37.2 |
| **UPF that is also HFSS (NPM 2018)** | 54.4 | 54.6 | 54.2 | 54.3 | 53.9 | 54.7 | 40.7 | 35.7 | 39.6 |

*Note:* HFSS = high in fat, salt or sugar; UPF = ultra-processed food; data are weighted to correct for non-random non-response of households; NPM = nutrient profiling model

**Table S2. The percentage of per capita daily foods, food energy in kcal, and food weight in grams consumed that was derived from foods that are high in fat, salt or sugar, ultra-processed foods, both and neither; UK National Diet and Nutrition Survey 2008/9-2018/19**

| | **All foods consumed; % (SD)** | **Food energy (kcal); % (SD)** | **Food weight (g); % (SD)** |
|---|---|---|---|
| **Neither UPFs nor HFSS (NPM 2004/5)** | 52.9 (14.4) | 31.7 (13.6) | 66.8 (18.6) |
| **Neither UPFs nor HFSS (NPM 2018)** | 54.1 (14.6) | 32.7 (13.9) | 66.0 (18.8) |
| **All UPF** | 33.6 (15.6) | 54.9 (15.7) | 29.7 (18.3) |
| **UPF only (NPM 2004/5)** | 15.0 (8.9) | 24.6 (10.5) | 18.2 (13.4) |
| **UPF only (NPM 2018)** | 15.8 (8.7) | 26.8 (11.3) | 17.8 (12.8) |
| **All HFSS (NPM 2004/5)** | 32.0 (10.6) | 43.7 (13.2) | 15.0 (10.9) |
| **All HFSS (NPM 2018**) | 30.0 (10.3) | 40.5 (12.7) | 16.2 (11.5) |
| **HFSS only (NPM 2004/5**) | 13.5 (8.1) | 13.4 (8.7) | 3.5 (4.3) |
| **HFSS only (NPM 2018)** | 12.3 (6.9) | 12.4 (8.0) | 4.3 (4.7) |
| **HFSS and UPF (NPM 2004/05)** | 18.5 (10.0) | 30.3 (13.8) | 11.5 (10.0) |
| **HFSS and UPF (NPM 2018)** | 17.7 (9.9) | 28.1 (12.9) | 11.9 (10.3) |
| **UPF that is also HFSS (NPM 2004/5)** | 55.1 (15.5) | 53.9 (17.3) | 40.0 (19.0) |
| **UPF that is also HFSS (NPM 2018)** | 52.3 (15.5) | 50.7 (17.3) | 40.2 (19.0) |

*Note.* HFSS = high in fat, salt or sugar (using the 2018 Nutrient Profiling Model); UPF = ultra-processed food; data are weighted to correct for non-random non-response of households.
